# Age specificity of cases and attack rate of novel coronavirus disease (COVID-19)

**DOI:** 10.1101/2020.03.09.20033142

**Authors:** Kenji Mizumoto, Ryosuke Omori, Hiroshi Nishiura

## Abstract

Age distribution of the cases with novel coronavirus disease (COVID-19) is rather different from that of influenza. In China, there were few reported cases among children [1] and serious or fatal child cases were also very infrequent. The age specificity is particularly important in designing details of social distancing, including school closure, as interventions, which is now recognized as the mainstream of interventions against COVID-19. Investigating the details of contacts, substantial susceptibility among children was demonstrated [2], but further insights into underlying mechanisms should be explored. Here we examined the age distribution of COVID-19 cases in Japan from January to March, 2020.

---

In Japan, a total of 313 domestically acquired cases have been confirmed as of 7 March 2020 (Figure 1). All these tested positive to reverse transcriptase polymerase chain reaction (RT-PCR), and they arose from suspected cases with close contact (n=2496). Male dominate confirmed cases (55.2%), and this finding is consistent with China [1]. Age category and gender were available for 173 male cases and 121 female cases. Out of 173 male cases, seven were aged 0-19 years, 84 were aged 20-59 years and 82 were aged 60 years and older, while three were aged 0-19 years, 69 were aged 20-59 years and 49 were aged 60 years and older for female. It is remarkable that there are very few child cases aged from 0-19 years in Japan.

**Figure 1.**
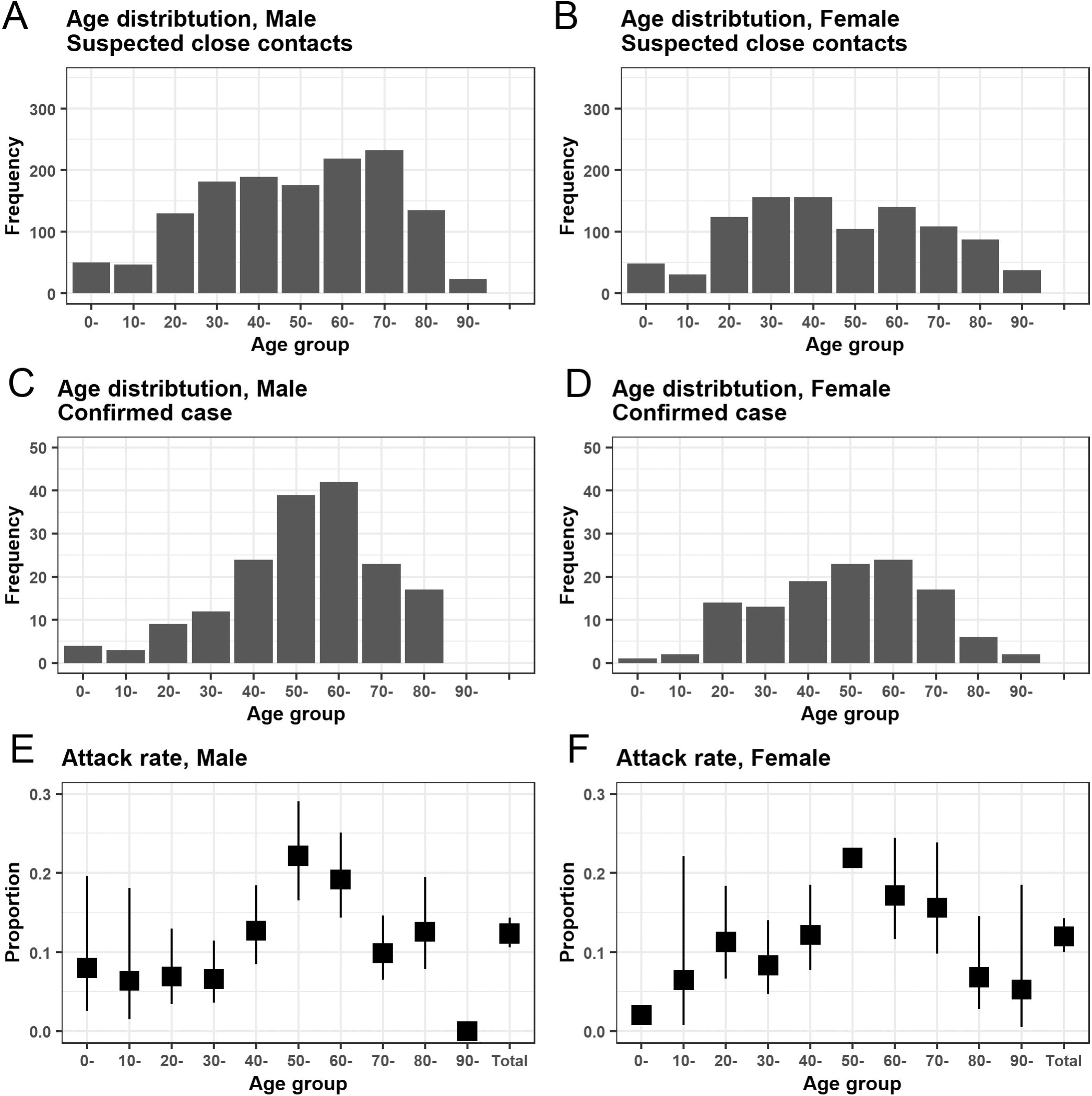
Age distributions of novel coronavirus disease (COVID-19) Age distributions of suspected individuals with close contact among male (A) and female (B), respectively. Age distributions of laboratory-confirmed cases among male (C) and female (D), respectively. Attack rate, i.e. cases out of suspected close contact, is shown by gender (male (E) and female (F)).

Because all sample data stems from exposed and suspected individuals, age-specific attack rate (AR) is calculable. Figures 1E and 1F show the AR estimates by age group and gender. AR was very low, 7.2% (95% Confidence Interval (CI): 3.0%, 14.3%) and 3.8% (95% CI: 0.8%, 10.6%), respectively, among male and female children aged from 0-19 years. The peak AR was seen in those aged from 50-59 years both for male, 22.2% (95% CI: 16.3%, 29.0%), and female, 21.9% (95% CI: 14.4%, 31.0%).

What our short analysis shows is that children are less likely to be diagnosed as cases, and moreover, the risk of disease given exposure among children appears to be low. Both the overall risk and the conditional risk of disease given exposure are likely to be the highest among adults aged from 50-69 years. The finding contradicts other widely circulating respiratory viral infections, e.g. seasonal influenza and respiratory syncytial virus infection, to which children are known to act as the focal host of transmission. How can the age-specificity happen? The most plausible explanation that we believe is immune imprinting to a similar virus among adults. Such virus must have continued to circulate in the human population by 20 years ago, and may be most intensely circulated by around 50 years ago. Severe acute respiratory syndrome-2 (SARS-2) coronavirus may be antigenically closely related to the old coronavirus, and infected adults in the present day may experience erroneous recognition of SARS-2 coronavirus. Although not focusing on the age specificity, a hypothetical discussion took place, suspecting of antibody dependent enhancement (ADE) as a potential biological mechanism of heterogeneous risks of death [3], which could explain the diagnosis of severe cases in higher age groups than in children, as seen in the present study.

In order to validate the hypothesis, it is ideal to conduct a seroepidemiological study which may involve multiple candidate antigens including other coronaviruses [4]. Examining the age-dependent strengths of immune cross reactivity, immune imprinting can be verified as the explanatory mechanism, which would have critical insights into minimization of unavoidable deaths.

## Data Availability

The datasets generated during and/or analysed during the current study are available from the corresponding author on reasonable request.

## Acknowledgments

H.N. received funding from the Japan Agency for Medical Research and Development (AMED) [grant number: JP18fk0108050]; the Japan Society for the Promotion of Science (JSPS) KAKENHI [grant numbers, H.N.: 17H04701, 17H05808, 18H04895 and 19H01074; K.M.: 18K17368], the Inamori Foundation, and the Japan Science and Technology Agency (JST) CREST program [grant number: JPMJCR1413].

## Authors’ contributions

All authors conceived the study and participated in the study design. K.M. collected and analyzed the data. H.N. and K.M. drafted the manuscript and R.O. and K.M. revised the the earlier versions of the manuscript. All authors edited the manuscript and approved the final version.

